# Strategy for identifying insulin pump use in Clinical Practice Research Datalink GOLD

**DOI:** 10.1101/2024.02.13.24302778

**Authors:** Rebecca Persson, Todd Sponholtz, Brenda N Baak, Susan S Jick

**Affiliations:** Boston Collaborative Drug Surveillance Program, Lexington, MA 02421; Boston University School of Public Health, Boston, MA

## Abstract

**Background:** Clinical Practice Research Datalink (CPRD) GOLD is an invaluable resource for clinical research. However, some exposures are difficult to capture, including continuous subcutaneous insulin infusion pump systems (“insulin pumps”). We present a strategy we developed to classify insulin pump users and to estimate the duration of pump use in CPRD GOLD. This was done to study adverse skin events in new adult pump users.

**Methods:** Insulin pump users were defined as patients who had a specific insulin pump code (prescription for an insulin pump cartridge or clinical code for continuous insulin infusion) in their record. Duration of use was defined as the continuous use of any insulin formulation commonly used in pump systems before and after the insulin pump specific code. Each patient’s pump start and end dates were calculated programmatically and then confirmed by manual review of the patient’s CPRD record.

**Results:** There were 1032 patients with an insulin pump specific code recorded in CPRD GOLD through December 2018, of which 302 met the inclusion criteria for our safety study. Due to high variability in the patterns of insulin use, programmatic determination of pump start and end dates was insufficient. The start and/or end dates of >50% of patients required adjustment upon manual review.

**Conclusions:** Insulin pump users in CPRD GOLD could be easily identified using this strategy, but we may have missed additional insulin pump users without specific pump codes. The duration of pump use, however, was difficult to capture. This strategy, though time intensive, is a useful tool for the study of insulin pumps.

**Funding:** This study was funded by AbbVie.

**Conflicts of Interest Statement:** Boston Collaborative Drug Surveillance Program (BCDSP) received funding from AbbVie for this study. Susan Jick and Rebecca Persson are employees of BCDSP. Todd Sponholtz and Brenda Baak were interns at BCDSP. Authors retain full and scientific control over the content of this manuscript.

**Disclosures:** This manuscript has not been peer-reviewed. We provide this information as a reference for other users of Clinical Practice Research Datalink GOLD.

This study is based in part on data from the Clinical Practice Research Datalink obtained under license from the UK Medicines and Healthcare products Regulatory Agency. The data is provided by patients and collected by the NHS as part of their care and support. The interpretation and conclusions contained in this study are those of the authors alone. This study was approved by the Independent Scientific Advisory Committee (ISAC) for Medicines and Healthcare products Regulatory Agency (protocol no: 19_216R).

## Introduction

The Clinical Practice Research Datalink (CPRD) GOLD, a large database of electronic medical records based in the United Kingdom, has been the source of data for research across many clinical areas.^1,2^ While most drugs are easily identified in the database, there are instances where the drugs, doses, or routes of administration can be difficult to capture.^3^ This difficulty arises when the drug is administered and/or prescribed in secondary care, such as by a consultant, resulting in gaps in the patient’s prescription record.^1^ This can pose problems for identifying drug devices such as subcutaneous insulin infusion pump systems (“insulin pumps”), and details of their use such as start and end dates.

We present a strategy used to identify new adult users of insulin pumps and the start and end dates of use in a study of adverse skin events in pump users in CPRD GOLD.^4^ We hope this information will be helpful for other researchers using these data for observational research of insulin pump use.

## Methods

CPRD GOLD is a large, prospectively collected, anonymized database encompassing over 500 UK general practices, covering over 10 million patients.^1,2^ It contains virtually complete electronic medical records and has been described extensively.^1,5-7^ Consultants and specialists (e.g., endocrinologists and diabetes clinics) are required to send letters describing relevant clinical events and diagnoses to the general practitioner (GP) whenever a patient is seen in secondary care. The contents of these letters are then entered into the GP’s computer records. When a patient is prescribed an insulin pump in secondary care, the GP will typically prescribe subsequent insulin prescriptions to be used in the pump. However, the GP may or may not code the insulin pump device in the GP record.

In order to account for the incomplete recording of insulin pumps and their start and end dates in CPRD GOLD, we developed the following strategy. From among all patients with an insulin prescription through December 2018, we identified patients with prescriptions for an insulin pump cartridge (Gemscript 31251021) or a code for a “subcutaneous infusion with insulin pump” (Read 7L10011), or “continuous subcutaneous infusion of insulin” (Read 7L10000) recorded with one of three insulin formulations commonly prescribed among patients with insulin pumps (Gemscript 50497020, 90309020, 89723020). We did not include patients with one of these three insulin formulations in the absence of a specific insulin pump code, as these formulations can also be used in injections. These patients comprised the cohort of insulin pump users. See Table.

**Table:**
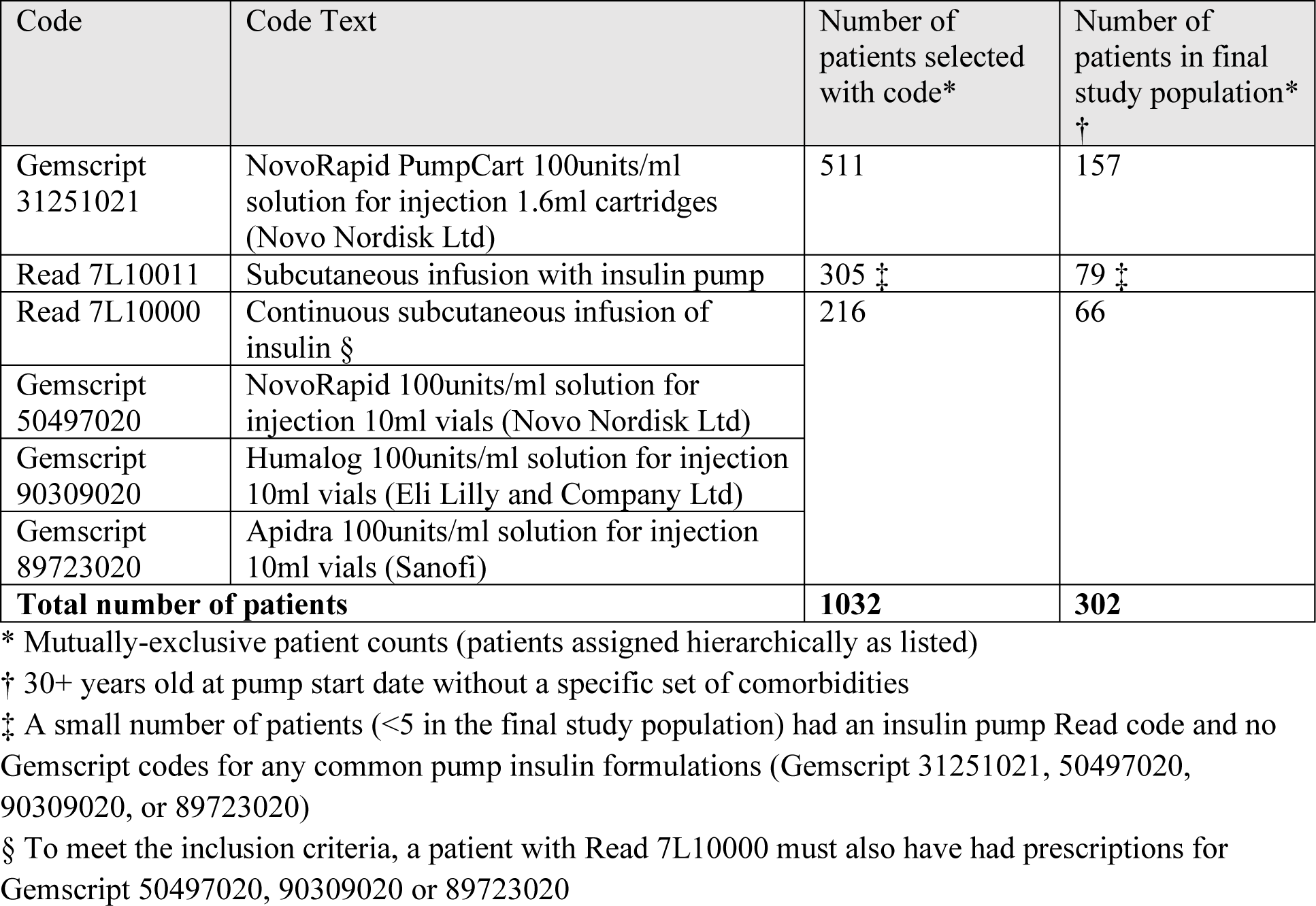
Codes and counts of insulin pump users in CPRD GOLD January 2019 release among all patients with an insulin prescription.

We determined that we could not depend on these codes alone to reliably estimate the start and end of pump use. The insulin pump cartridge Gemscript code was first used in October 2014. However, many patients with this code had continuous use of the other insulins commonly used in pump systems before that date. Similarly, some patients with a Read code for continuous insulin infusion had continuous use of one of the three insulin prescriptions before and after the continuous infusion Read code. Thus, we programmatically assigned tentative dates for the start and end of pump use based on continuous use of one of the four codes (three insulin codes plus the pump code) commonly used in pump systems as follows.

The first recording of an insulin pump cartridge or continuous insulin infusion was assigned to each patient as the tentative pump start date. If a patient had continuous use (defined below) of another insulin commonly used in pump systems for more than 30 days before the tentative start date, we assigned a final pump start date to the beginning of the continuous use of these insulin prescriptions. Otherwise, we used the tentative start date as the final date. Similarly, we assigned a final pump end date to the end of the continuous use of insulin prescriptions used in pump systems when the record contained no new prescriptions for any pump insulins or diagnoses for insulin pump use after the end of filled use + a 90-day washout in the absence of eligible “bridging” (defined below). Each final pump start and end date was manually reviewed and confirmed with consensus of up to 4 researchers. New episodes of insulin pump use recorded after the final pump end date were not assessed in our study but the method described above could be repeated to capture subsequent periods of insulin pump use.

We employed a number of assumptions to define continuous insulin pump use and calculate person time of exposure. In our final study population, the median number of days between insulin prescriptions was 9.6 days/vial. Thus, we assigned 10 days of filled use per insulin vial. For patients with a Read code for insulin pump without a corresponding Gemscript insulin prescription, we assigned 30 days of filled use, assuming the patient received their first insulin prescription from a specialist. Lastly, as patients with type 1 diabetes must use insulin, we assumed that any patient with an extended period with no insulin prescription records must have received additional prescriptions from a specialist. For patients with continuous pump use with the same insulin formulation before and after a period with no insulin records, we assumed the patient was using the pump throughout and “bridged” the gap in prescriptions with prescriptions from another clinician not recorded in CPRD GOLD. Patients were defined as having continuous use in the presence of concomitant use of non-pump insulins (e.g., insulin pens) as long as the filled use of pump insulins was continuous. (See Figure.)

**Figure Legend:**
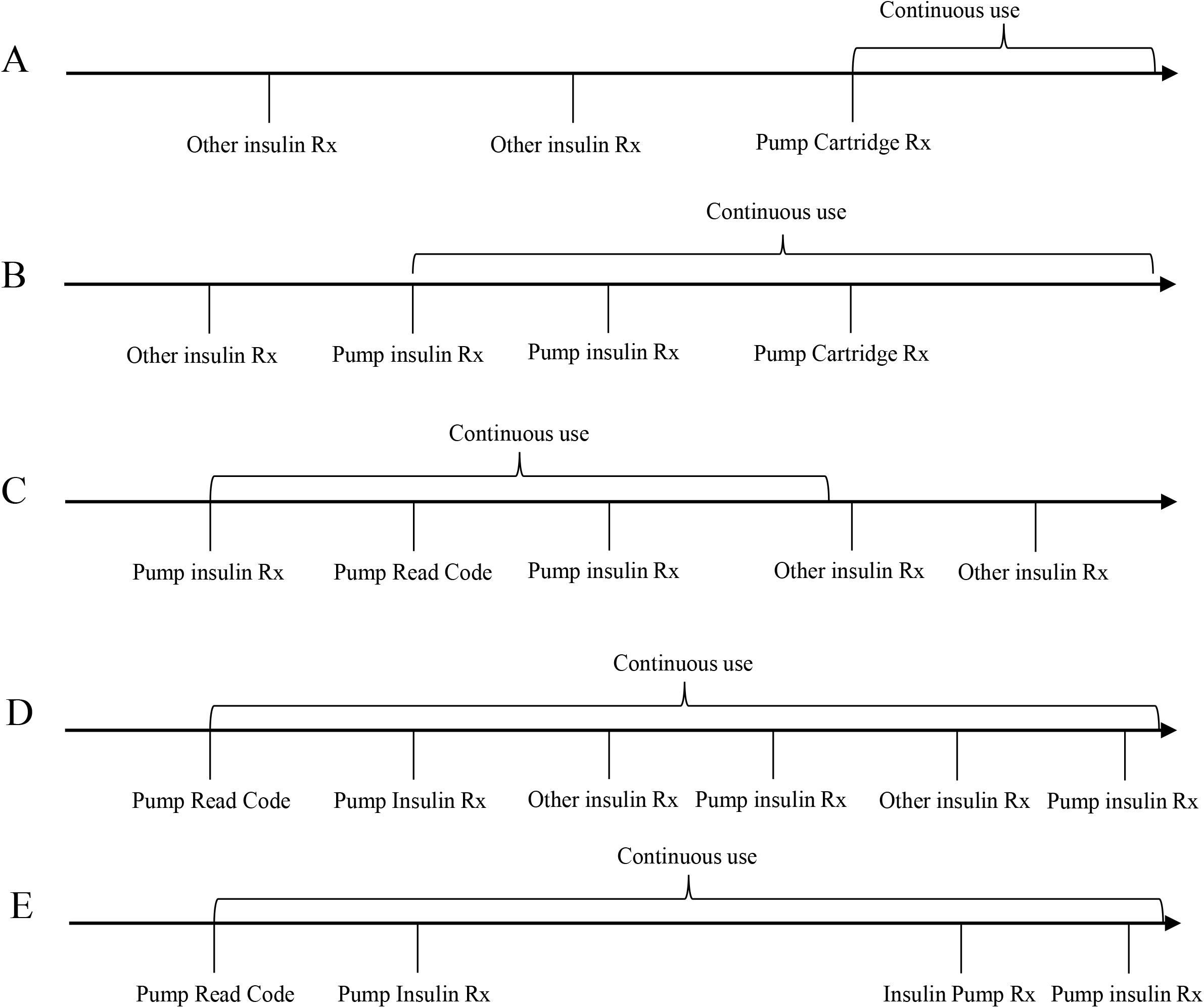
Assignment of continuous insulin pump use in various patient scenarios. (A) Patient had insulin pump cartridge prescription (Gemscript 31251021) and no prior pump related codes; (B) Patient had regular use of insulins used in pump systems (Gemscript 50497020, 90309020, or 89723020) before their insulin pump cartridge prescription; (C) Patient had regular use of insulins used in pumps systems before and after a Read code for continuous insulin infusion (Read 7L10011 or 7L10000) and then switches to other non-pump insulin formulations; (D) Patient had regular use of insulins used in pump systems with concomitant use of other insulins; (E) Patient had an extended period with no insulin prescriptions with regular use of pump insulins before and after the gap. Rx = prescription.

## Results

There were 1032 (0.5%) patients who met our insulin pump definition among 176,870 patients with an insulin prescription or a continuous insulin infusion Read code. Approximately half of pump users were identified through a prescription for an insulin pump cartridge (Gemscript 31251021). (See Table.) The majority were female (N=614, 60%) and the median age at first specific pump code was 29 years.

Only 302 patients met the criteria to be included in our safety study (age 30+ at pump start date and without a selected set of study-specific comorbidities),^4^ thus our strategy for determining duration of use was based on a subset of insulin pump users. Upon manual review, we adjusted the tentative start and/or end dates of more than 50% of patients, as the programmed algorithm was overly sensitive to occasional use of insulin pens or other injectable insulins as well as to inconsistent spacing of insulins used in pump systems throughout the calendar year. Most patients (N=272; 90%) had continuous insulin pump use through the end of the record and 30 patients (10%) discontinued pump use after a median 18 months. There were 10 (3%) patients who had a gap in continuous insulin pump use of 6 to 18 months with no recorded insulin prescriptions that were “bridged” by the continuous use algorithm.

## Discussion

This strategy was useful for studying use of insulin pumps in CPRD GOLD, however, there was considerable variability in the pattern of insulin use between patients, and the automated determination of duration of use was insufficient. Although time-intensive, manual review was required to assign plausible pump start and end dates.

Although we did not conduct a formal validation study, this strategy was designed to be specific (i.e., minimize the number of non-pump users included in the case definition). It is, however, likely to have low sensitivity (i.e., there may be many additional insulin pump users whose CPRD GOLD records do not clearly designate pump use). The most useful pump specific code (Gemscript 31251021) was introduced in October 2014. It is possible that a study conducted in data from 2015 and later may have a higher sensitivity. Furthermore, as our safety study population was limited to adult insulin pump users,^4^ we did not account for differences in coding or patterns of use in pediatric patients.

This strategy may result in some misclassification of insulin pump exposed person-time, particularly at the start and end of use as well as during any extended periods with no recorded insulin prescriptions. Researchers using this strategy should consider conducting sensitivity analyses to assess the potential effects of misclassified person-time.

CPRD GOLD is an invaluable resource for research. However, some exposures, including insulin pump use, are difficult to capture. This strategy is a useful tool for studying the use of insulin pumps in CPRD GOLD.

## Data Availability

Per their license agreement with CPRD, the authors cannot make data available. Requests to access data can be made directly to the CPRD.

